# Evaluation of RNA extraction free method for detection of SARS-COV-2 in salivary samples for mass screening for COVID-19

**DOI:** 10.1101/2021.03.15.21253570

**Authors:** Sally A. Mahmoud, Esra Ibrahim, Subhashini Ganesan, Bhagyashree Thakre, Juliet G Teddy, Preeti Raheja, Walid Z Abbas

## Abstract

In this current COVID - 19 pandemic, there is a dire need for cost effective and less time-consuming alternatives for SARS-COV-2 testing. The RNA extraction free method for detecting SARS-COV-2 in saliva is a promising option, this study found that it has high sensitivity (85.34%), specificity (95.04%) and was comparable to the gold standard nasopharyngeal swab. The method showed good percentage of agreement (kappa coefficient) 0.797 between salivary and NPS samples. However, there are variations in the sensitivity and specificity based on the RT-PCR kit used. The Thermo Fischer-Applied biosystems showed high sensitivity, PPV and NPV but also showed higher percentage of invalid reports. Whereas the BGI kit showed high specificity, better agreement (kappa coefficient) between the results of saliva and NPS samples and higher correlation between the Ct values of saliva and NPS samples. Thus, the RNA extraction free method for salivary sample serves as an effective alternative for SARS-CoV 2-testing.

## Introduction

The current pandemic caused by SARS-COV-2 virus, widely known as COVID-19, has affected more than 75 million people worldwide and as of 20^th^ December 2020 has caused more than 1.6 million deaths according to WHO [1]. Early identification, isolation and quarantine of contacts play a vital role in controlling the pandemic. The high number of COVID-19 cases and fast spreading ability of the virus warrants mass screening which involves large number of diagnostic tests being conducted. Currently sample collection with nasopharyngeal swabbing followed by RT-PCR of is the gold standard for diagnosis of COVID-19 [2].

Nasopharyngeal swabs requires trained personnel to collect the sample, there is a risk of exposure for the healthcare personnel procuring the sample, discomfort to the patients during the procedure, there are also contraindications for doing nasopharyngeal swabbing like coagulopathies, anti-coagulant therapy and deviated nasal septum. Moreover, when the swabbing is not performed correctly, adequate sample might not be obtained, leading to inconclusive/invalid results and the need to repeat swabbing. This method of sample collection also requires the use of PPE by the healthcare personnel collecting the swab, as there is a high risk of exposure, leading to additional drain on resources. All the above-mentioned aspects suggest the need for alternative methods of testing and diagnosis for COVID-19 and saliva seems to be an excellent alternative available.

The human saliva has hundreds of microbial species, many of which are related to diseases affecting humans [3],including hereditary, autoimmune diseases, malignancies and infections [4,5]. Saliva has also been used as an important diagnostic tool in previous coronavirus infections like severe acute respiratory syndrome (SARS) and Middle East respiratory syndrome (MERS) [6].

Diagnosis of viral infections from salivary samples depends on various factors like the presence of the virus in the saliva and the load of viral particles like DNA, RNA, antigens, host antibodies, the duration and stage of infection as some viruses can be detected in saliva almost a month after infection [7,8].

In the context of COVID-19, the saliva plays a crucial role in the transmission of the disease, as droplets are the main source of human to human transmission. The saliva contains the viral particles, anti-SARS-COV-2 antibodies, infected host cells, which can be used for diagnostic as well as prognostic purposes. [9,10] Further in COVID-19, the infection of salivary glands occurs earlier in the disease [11], the viral load in saliva is highest during the first week of onset and it can be detected in saliva as long as 25 days after the onset of infection, all this suggests saliva as an effective alternative non-invasive method for diagnosis of COVID-19 [12].

Saliva testing also offers additional advantages during mass testing, as it is non – invasive, does not require a trained healthcare personnel to collect the sample, therefore alleviates the need for PPE, swabs and the issues of demand of PPE and other resources, no risk of exposure for the health personnel as the patient themselves can collect the sample, there are no contraindications for this method of sample collection, its more comfortable for patients especially when multiple testing are done and using salivary samples disease monitoring can also be done[11-13].

Hence this study aims to explore how efficient is RNA extraction free method for detection of SARS-COV-2 in saliva, as an alternative specimen for COVID-19 testing.

### Objectives

1. To compare the sensitivity and specificity of RNA extraction free method for molecular testing of SARS-COV-2 of salivary sample vs nasopharyngeal samples
2. To compare the efficiency of RNA extraction free method of salivary sample in established PCR-based testing with BGI Genomics’ 2019-nCoV Fluorescence Detection Real-Time RT-PCR Kit and Thermo fischer/ Applied Biosystems TaqPath COVID-19 CE-IVD RT-PCR Kit

## Methodology

### Sample size calculation

The SARS-COV-2 infection positivity was around 5% in our lab, at the time of the study. With the recommendation based on Bujang et al [14] we calculated the minimum sample size to be 380 participants, including 19 positive cases to achieve a minimum power of at least 80%, at a significance level of 5%, to detect a sensitivity of 95%. This sample would also be able to detect a specificity of 95%.

### Methods

The study was approved by the Department of Health (DOH) Institutional review board (IRB), Abu Dhabi. The study followed all regulations and guidelines of the IRB. The participants of the study were well-informed about the details of the study and informed consent was obtained. After obtaining the consent from the participants, one nasopharyngeal swab (NPS) and salivary sample was collected from each participant. Totally 600 people participated in the study. The NPS samples were collected by trained healthcare personnel as per protocol in place and were immediately transported to the lab. The salivary samples were collected in a sterile Dnase/Rnase free container without any stabilizing agent. Instructions were given to all the participants before saliva collection. The preconditions for saliva collection include no food, drinks, smoke nor oral hygiene products in the last one hour prior to sample collection. Participants were asked to pool saliva in their mouth for 1–2 minutes. The collected salivary samples were transported to the lab, in temperature-controlled boxes at 2-8 degrees Celsius, where they were processed by a nucleic acid extraction free method for SARS-COV-2 detection.

### RNA extraction free direct method

It is a nucleic acid extraction free method for SARS-COV-2 detection following the protocol of Saliva-Direct test, a saliva-based, nucleic-acid-extraction-free, qRT-PCR method for SARS-CoV-2 detection [15]. The Saliva sample collected is first treated with proteinase K followed by a heat inactivation step and is then directly used as input in the RT-qPCR test using validated primer and probe sets.[Figure1] The RT-PCR was performed using BGI Genomics’ 2019-nCoV Fluorescence Detection Real-Time RT-PCR Kit and Thermo fisher/ Applied biosystems kit.

**Figure 1:**
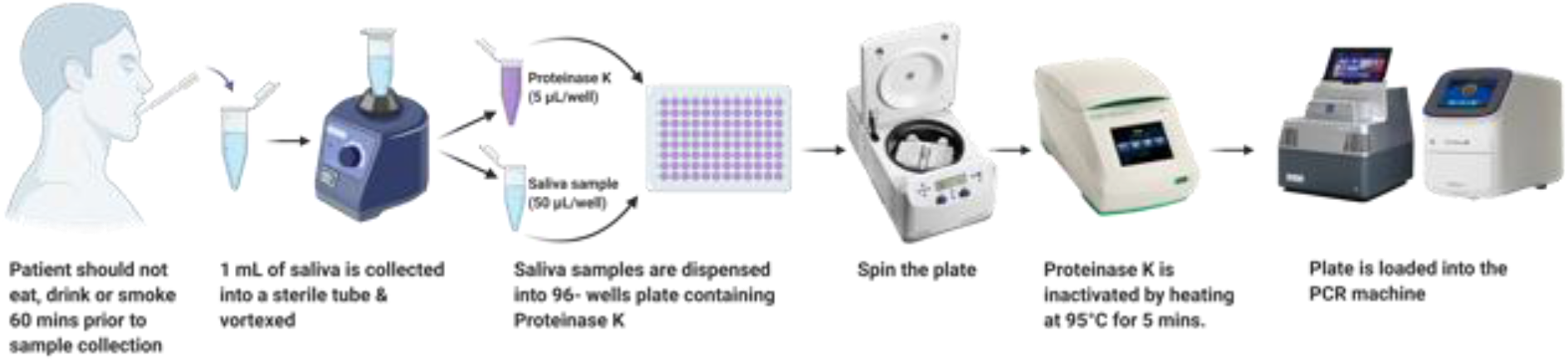
Process of RNA extraction free method for molecular testing of SARS-COV-2.

### The BGI Genomics’ 2019-nCoV Fluorescence Detection Real-Time RT-PCR Kit [BGI kit] [16]

The Real-Time Fluorescent RT-PCR Kit for Detecting SARS-2019-nCoV are used for the identification of SARS-CoV-2 RNA in nasopharyngeal swabs, throat swabs and Broncho alveolar Lavage Fluid (BALF) from patients with SARS-CoV-2.

The qPCR procedure was performed following manufacturer’s instructions and was optimized for processing saliva samples. For optimization, results with high CT values for viral genes (ORF1AB) at FAM channel (CT value > 35 and ≤38) or with abnormal amplification curves were reprocessed for confirmation. Samples with CT values >38 and S shaped amplification curve are considered in the Grey zone and were reprocessed for confirmation. If the same result was obtained with the repeat testing, the result was confirmed as positive. This optimization was done to avoid false positives results at higher CT values.

### The Thermo fischer/ Applied Biosystems TaqPath COVID-19 CE-IVD RT-PCR Kit [Thermo fisher kit][17]

TaqPath COVID-19 CE-IVD RT-PCR Kit contains the reagents and controls for real-time reverse transcription polymerase chain reaction (RT-PCR) test intended for the qualitative detection of nucleic acid from SARS-CoV-2 in upper respiratory specimens (such as nasopharyngeal, oropharyngeal, nasal, and mid-turbinate swabs, and nasopharyngeal aspirate) and bronchoalveolar lavage (BAL) specimens from individuals suspected of COVID-19. It is a multiplex assay that contain three primer/probe sets specific to different SARS-CoV-2 genomic regions and primers/probes for bacteriophage MS2. The manufacturer instructions were followed, and reporting was done as per the instructions.

To compare the efficiency of RNA extraction free method of salivary sample in RT-PCR based testing with BGI RT-PCR kit and Thermo fischer/ Applied Biosystems RT-PCR Kit, 258 random samples out of these 600 salivary and NPS were processed through the RT-PCR machine using the Thermo fischer/ Applied Biosystems kit. The reports of the salivary sample processed through both these kits were compared with the results of the gold standard NPS results.

## Results

600 salivary samples were included in our study and 5 of these 600 (0.8%) salivary samples turned invalid. The results of the salivary sample were compared with the NPS sample results. [Table 1]

**Table 1:** Shows the results of salivary sample compared to gold standard NPS results using the BGI RT-PCR kit (n=600)

| NPS sample [BGI KIT] |  | Positive | Negative | Total |
| --- | --- | --- | --- | --- |
| RNA extraction free method for saliva (BGI kit) | Positive | 198 | 18 | 216 |
|  | Negative | 34 | 345 | 379 |
|  | Total | 232 | 363 | 595 |

The sensitivity detected was 85.34%, specificity – 95.04%, Positive predictive value (PPV) – 91.67%, positive predictive value (NPV)-91.03%, the measurement agreement - Kappa coefficient was 0.797 (p<0.001) when compared to the gold standard NPS results. The mean Ct value for saliva samples was 11.29 ± 15.21 and the mean for NPS samples Ct value was 10.93 ± 13.55. The difference was not found to be statistically significant (p value – 0.433). The correlation between the Ct values of saliva and NPS were found to be 0.692 with a significant p value <0.001

Out of these 600 samples 258 random samples were processed for RT-PCR by RNA extraction free method using the Thermo fischer/ Applied Biosystems kit, of which 27 salivary samples turned to be invalid. The results of the salivary sample were compared with the gold standard NPS sample. [Table 2]

**Table 2:**
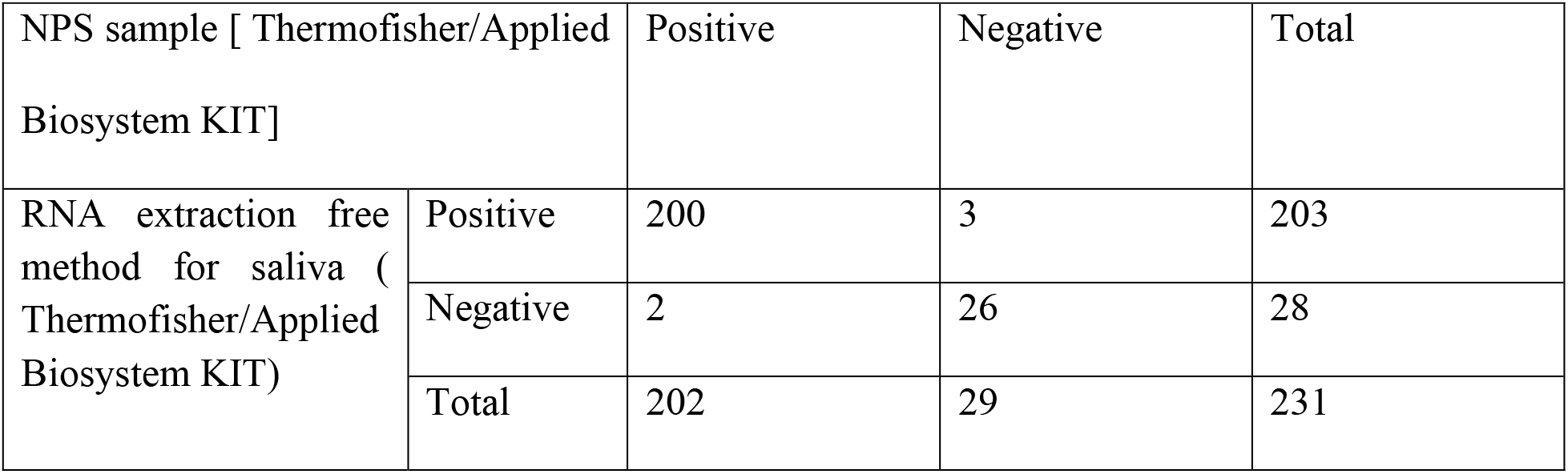
Shows the results of salivary sample compared to gold standard NPS results using the Thermo fisher RT-PCR kit (n=258)

For comparison, matched 258 samples that were processed with both the BGI RT-PCR Kit and Thermo fischer/ Applied Biosystems kit were used for further analysis, in this 4 samples turned to be invalid. Table 3 shows the results of the matched 258 salivary and NPS samples using the BGI kit [Table 3]

**Table 3:** Shows the results of the 258 matched salivary sample processed through the BGI kit compared to gold standard NPS results (n=258)

| NPS sample [BGI KIT] |  | Positive | Negative | Total |
| --- | --- | --- | --- | --- |
| RNA extraction free method for saliva (BGI kit) | Positive | 109 | 10 | 119 |
|  | Negative | 19 | 116 | 135 |
|  | Total | 128 | 126 | 254 |

The sensitivity, specificity, PPV, NPV and the percentage of agreement (kappa agreement %) between the two different kits were calculated and compared. [Figure 2]. There was statistically significant difference between the two kits in terms of sensitivity, PPV, agreement percentage and the percentage of invalid results.[Table 4]

**Table 4:**
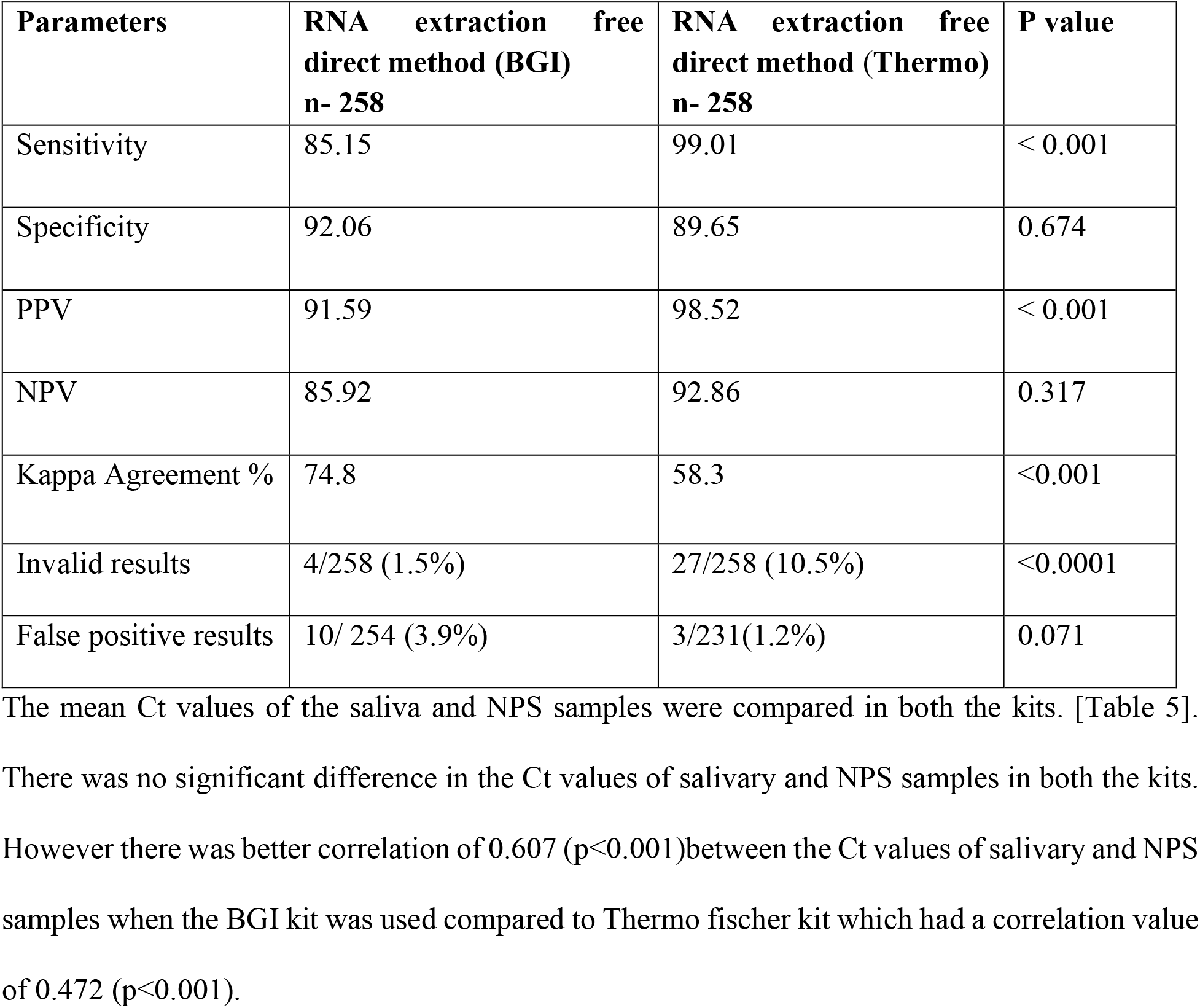
Compares the parameters between the BGI and Thermo fisher kit.

| Parameters | RNA extraction free<br>direct method (BGI)<br>n- 258 | RNA extraction free<br>direct method (Thermo)<br>n- 258 | P value |
| --- | --- | --- | --- |
| Sensitivity | 85.15 | 99.01 | < 0.001 |
| Specificity | 92.06 | 89.65 | 0.674 |
| PPV | 91.59 | 98.52 | < 0.001 |
| NPV | 85.92 | 92.86 | 0.317 |
| Kappa Agreement % | 74.8 | 58.3 | <0.001 |
| Invalid results | 4/258 (1.5%) | 27/258 (10.5%) | <0.0001 |
| False positive results | 10/ 254 (3.9%) | 3/231(1.2%) | 0.071 |

**Figure 2:**
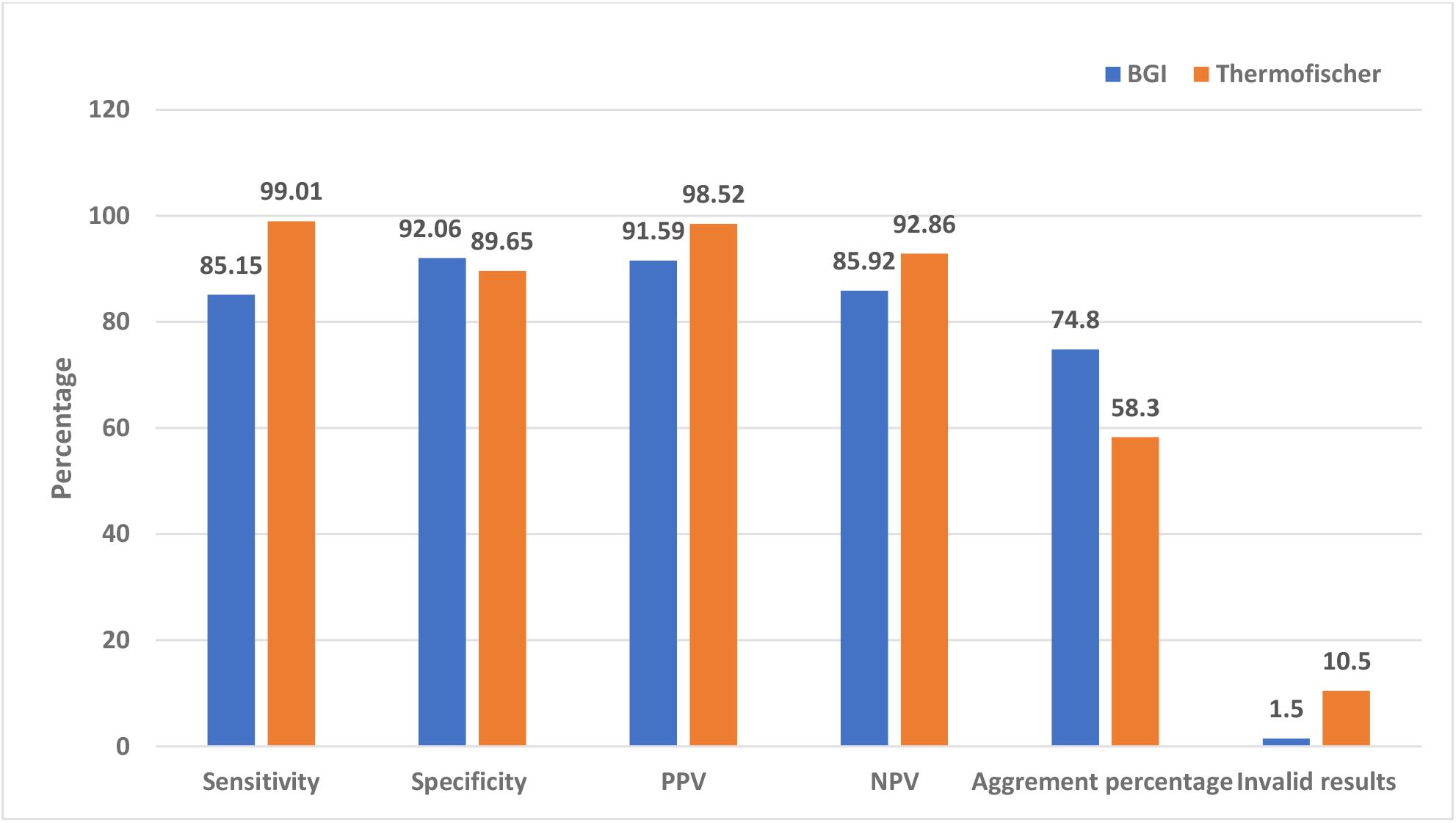
Graph comparing the various parameters between the BGI and Thermo fisher kit.

The mean Ct values of the saliva and NPS samples were compared in both the kits. [Table 5]. There was no significant difference in the Ct values of salivary and NPS samples in both the kits. However there was better correlation of 0.607 (p<0.001)between the Ct values of salivary and NPS samples when the BGI kit was used compared to Thermo fischer kit which had a correlation value of 0.472 (p<0.001).

**Table 5:** Compares Ct values between the saliva and NPS samples.

| Method | Sample | Mean ± SD | *p* value | Correlation | *p* value |
| --- | --- | --- | --- | --- | --- |
| **BGI Kit** | Saliva | 14.19 ± 15.66 | 0.632 | 0.607 | <0.001 |
| | NPS | 13.80 ± 13.43 | | | |
| **Thermo**<br>**Fisher Kit** | Saliva | 20.52 ± 12.79 | 0.343 | 0.472 | <0.001 |
| | NPS | 21.24 ± 10.05 | | | |

## Discussion

The study showed that the saliva RT-PCR (RNA extraction free method) has high sensitivity and specificity and is comparable to the gold standard nasopharyngeal swab (NPS). Studies reported earlier have shown similar results, where the sensitivity of saliva samples in detecting SARS-COV-2 ranges from 73% to 91% and the specificity ranges from 97 % to 98.9% [18,19,20].

In our study the PPV (91.6%) and NPV (91.03%) was high for direct (RNA extraction free method) saliva testing and similar results have been reported in other studies also [18,20].

The agreement between the results of saliva and NPS samples was good based on kappa coefficient. The values were similar to other studies that have demonstrated 0.68 to 0.85 kappa coefficient on agreement of salivary samples with the nasopharyngeal swabs [18,21]. Thus, this study demonstrates that even with the RNA extraction free method the results agreement was high between salivary and NPS samples.

However, in this study we observed that there were variations in the sensitivity and specificity in the direct (RNA extraction free method) method based on the kit used. The Thermo Fischer-Applied biosystems showed high sensitivity, PPV and NPV, whereas the BGI kit showed high specificity and better agreement (kappa coefficient) between the results of saliva and NPS samples.

In the BGI kit the mean Ct values of both saliva and NPS was low compared to the mean Ct values of saliva and NPS samples when Thermo fisher Applied Biosystem Kit was used. But both the kits did not show any statistically significant difference in Ct values between the saliva and the NPS samples. Similarly, studies have reported that there was no significant difference in the Ct values of saliva and the NPS samples [22]. There are studies that have also reported significant difference in the Ct values of saliva and NPS samples [23].The differences reported might be because earlier studies have also showed that the saliva viral load declines more rapidly than NPS viral load after initial two weeks of infection [24] which might have played a role in the significant difference in the Ct values.

This study also observed significant correlation between the Ct values of saliva and NPS samples and the correlation was better observed with the BGI kit, similar significant correlations were reported in other studies [25].

From the results obtained, our study found that the drawback of the Applied Biosystems kit was the number of invalid reports, which was significantly higher than the BGI kit. Which might be because it is a multiplex assay and hence the presence of impurities or inhibitors in body fluids interferes with the assay [26].

In the RNA extraction free method 3.9 % and 1.2% of the patients with a negative NPS had a positive saliva sample result using the BGI kit and the Thermofisher Applied Biosystems kit respectively. Other studies have also reported similar results where saliva samples showed higher detection rates at controlled conditions [21,27]. This variation in results might be due to preanalytical factors affecting the quality of the NPS samples used. The preanalytical factors include the skill of the trained healthcare professional, the technique used, transport media, temperature control and storage conditions[13]. The effect of these pre-analytical factors are less pronounced in the saliva sample if 1-hour fasting is strictly applied.

Hence the study shows that saliva can be a reliable alternative for SARS-COV-2 testing and similar studies have shown saliva to be a reliable sample even without the RNA extraction process.[28]. As the sensitivity and the PPV of the saliva sample is high and comparable to NPS sample, this method can be highly useful in identifying patients early and isolating them to limit spread.

During the current pandemic where mass testing is mandated, the RNA extraction free method of saliva sample is time saving and it precludes the need for specific collection devices, transport media and skilled personnel. This can considerably reduce the burden on the health care industry and staff. Moreover, when manufacturing and transport of goods have taken a hard hit in the current crisis, these simple techniques enable testing of low volume and minimally processed sample and is not affected by the supply chain constraints[15].

### Strengths and Limitations

The strength of this study is that it not only evaluates the RNA extraction free method for molecular testing of SARS-COV-2 of salivary sample, but also validates and compares different RT – PCR kits for extraction free method of saliva sample. Not many studies have evaluated these.

Limitation of this study is that the study did not consider the number of days the patient has been infected or symptomatic in analysis, as the viral load in the saliva varies with the stages of infection which might have affected the results of the study.

## Conclusion

The study concludes that the RNA extraction free method for molecular testing of SARS-COV-2 in salivary sample serves as an effective alternative for COVID-19 testing. We like to emphasize on the fact that the nature of salivary sample collected, the method of extraction, RT-PCR kit used can all influence the results. Yet saliva collection is simple, no specialized devices needed for collection, the extraction free method saves time, effort and cost that enables mass testing and quick and early reporting to control spread. Thus, despite the limitation’s RNA extraction free method for molecular testing of SARS-COV-2 in salivary sample can be broadly implemented as an alternative for SARS-COV-2 detection.

## Data Availability

The data is available with the author, Dr. Sally, Director of Biogenix G42 lab and will be produced on request

## Declarations

### Ethics approval and consent to participate

The Ethics approval was obtained from Department of Health (DOH) Institutional review board (IRB), Abu Dhabi

All methods were carried out in accordance with relevant guidelines and regulations. The Department Of Health (DOH) Institutional review board (IRB), Abu Dhabi

Consent for publication – written informed consent were obtained from all the participants

## Acknowledgments

We acknowledge all lab personnel, skilled technicians who provided help during the research and preparation of the manuscript

